# Contralateral spread of unilateral tremor in Parkinson’s Disease

**DOI:** 10.1101/2025.02.20.25322608

**Authors:** Jacopo Pasquini, Nicola Pavese, Roberto Ceravolo, Rick C. Helmich, Günther Deuschl

## Abstract

**Background:** Parkinsonian tremor usually starts unilaterally. The mid-term prognosis of this lateralized tremor is unknown, as is the development of tremor in the contralateral arm.

**Objective:** To investigate the emergence of contralateral tremor in the Parkinson-Progression-Marker-Initiative database, with data available for 7 years.

*Methods:* Tremor Amenable for surgery (TAS) was defined as any rest, postural or kinetic tremor with amplitude >1 cm (MDS-UPDRS score ≥2) as this criterion is commonly accepted for inclusion in surgical studies. Tremor was analyzed by side mainly in the off-medication state.

*Results:* At baseline, 348 (87.7%) of the 397 patients with Parkinson’s disease had tremor at least on one side of the body. 183 (46%) had only mild tremors but 165 (41.6%) had TAS. 159 patients (40.1%) had lateralized TAS and 6 (1.6%) had bilateral TAS. Among patients with unilateral TAS, 40 patients (25.8%) developed contralateral TAS at 3 years, 49 patients (30.8%) at 5 years, and 61 patients (39%) at 7 years. The side more affected by tremor was also more affected by other cardinal symptoms. In 159 patients with initially unilateral TAS, tremor severity did not increase on the tremor-dominant side over the 7-year period. However, there was an increase in tremor on the contralateral side. This was associated with a clear increase in bradykinesia and rigidity on both sides.

**Conclusion:** The study findings may prove beneficial in counselling patients with TAS, and may also provide an explanation as to why the worsening of tremor is not correlated with overall disease progression.

## Introduction

Tremor is the eponymous symptom of the shaking palsy and is yet poorly understood in many respects. Particularly, the long-term course of this troublesome and disabling symptom has only been studied for relatively short periods.^1^ A distinctive feature is the unilateral onset of PD,^2^ and particularly the classical resting tremor, which can facilitate the neurologist’s overall clinical diagnosis. Unilateral tremor may progress to the contralateral side but evidence regarding the timing and severity of this phenomenon is lacking. With the focus on early symptoms of PD, studies on the natural course are now increasingly popular, but their focus has only rarely been on tremor.^3, 4^ Particularly, the question for the ‘time-to-contralateral tremor’ has to our knowledge never been studied. In the context of tremor treatment, this is becoming an increasingly pertinent question, given the availability of invasive treatments for PD symptoms on a single body side, such as Magnetic Resonance guided Focused Ultrasound (MRgFUS) and in some cases deep brain stimulation. Indeed, it is not known if and when the second side will be affected in patients with unilateral tremor. Here we address this question by analyzing the cohort of patients from the Parkinson Progression Marker Initiative^s^ who have been assessed prospectively on an annual basis using the MDS-UPDRS. The main research question is when and to what extent functionally impairing tremor develops on the second side after a relevant tremor has been diagnosed on the first side. Using the anchors defined by the MDS-UPDRS, we define a tremor severity >1 cm at the fingertips under resting or postural or kinetic conditions as ‘tremor amenable to surgery’ (TAS).

## Methods

### Participants

This paper uses the clinical database of the Parkinson Progression Marker Initiative (PPMI).^s^ The data includes demographic and clinical characteristics of 397 sporadic PD participants, also included in a previous analysis from our group.^4^ This analysis used data from the PPMI cohort enrolled between 2010-2020. Protocol information for The Parkinson’s Progression Markers Initiative (PPMI) 001 AM 12 can be found at the following link: https://www.ppmi-info.org/study-design/archive-of-research-docs-and-sops.html. The PPMI was designed to be an eight-year natural history study (with a minimum of five-year involvement) of de novo idiopathic PD participants. All PD subjects were planned to have an annual assessment of the motor exam in a practically defined off-state and a repeat on-state assessment one hour after receiving their usual PD medication in clinic. For this study, we considered the first seven years of follow up after enrollment.

Since not all patients attended their assessment each year, in **supplementary Table 1** we show the number of participants that attended each follow up.

The clinical data of the cohort at baseline are summarized in Table 1.

**Table 1.** Baseline clinical characteristics of the PPMI cohort included in this study.

|  | **Baseline characteristics** |
| --- | --- |
| **Number of eligible participants** | 397 |
| **Males/Females** | 261/136 (66%/34%) |
| **Age at diagnosis (years), mean (SD), range** | 61.91 (9.59), 34-85 |
| **Disease duration at enrollment (months), mean (SD), range** | 6.76 (4.07), 0-37 |
| **MDS-UPDRS III, mean (SD), range** | 21.00 (8.88), 4-51 |
| **Bradykinesia score<sup>1</sup>, mean (SD)** | 8.32 (4.90) |
| **Rigidity score<sup>2</sup>, mean (SD)** | 3.82 (2.66) |
| **Hoehn & Yahr, median (range)** | 2 (1-2) |

<sup>1</sup>Sum of MDS-UPDRS of subitems of items 3.4, 3.5, 3.6, 3.7, 3.8; <sup>2</sup>Sum of subitems of item 3.3; Abbreviations: MDS-UPDRS III = Movement Disorders Society – Unified Parkinson Disease Rating Scale part III; SD = standard deviation.

### Definition of tremor amenable for surgery (TAS)

Within the PPMI database, tremor is measured with the MDS-UPDRS. Lateralized items related to tremor come only from the physician exam of part III (motor score) covering information for postural tremor (Item 3.15), kinetic tremor (3.16), and rest tremor (3.17) for the upper extremity, and for the lower extremity for rest tremor only (3.17). All these tremor manifestations are measured with the following anchors documented as the maximal amplitude during the defined exams. 0: No tremor. 1: Slight: < 1 cm. 2: Mild: > 1 cm but < 3 cm. 3: Moderate: 3 - 10 cm. 4: Severe: > 10 cm.

The further tremor items 3.17e (rest tremor amplitude-lip/jaw) and 3.18 (constancy of rest tremor) are excluded from this analysis as they cannot contribute to the definition of lateralized tremor.

For the purpose of this study, we define a tremor possibly eligible for invasive treatments as having an amplitude of more than 1 cm at fingertips (corresponding to the anchor score of 2 in the upper limbs tremor items) and call this a ‘tremor amenable for surgery (TAS)’. Such TAS is assumed if at least one of the upper limbs tremor scores for rest, postural and kinetic tremor is rated as 2 or more. This criterion is commonly used as an inclusion criterion for studies testing invasive interventions such as MRgFUS.^6^

### Definition of lateralized TAS

The TAS criterion allows the identification of participants with a lateralized TAS (i.e. an upper limb tremor subitem score 2: 2, with contralateral upper limbs subitems scores ≤ 1), a bilateral TAS (an upper limb tremor subitem score 2: 2 both on right and left sides), and participants without TAS (all upper limbs tremor subitems scores ≤ 1).

### Definition of lateralized scores

A total lateralized tremor score was also calculated by summing the lateralized MDS-UPDRS 3.15, 3.16 and 3.17 subitems. Here, we also included the rest tremor lower limb score in the sum, as this contributes to the overall burden of tremor score on each side. Therefore, the lateralized tremor score ranges between 0 and 16 points. Additionally, for comparison, we have calculated the lateralized scores for bradykinesia and rigidity. The lateralized bradykinesia score was calculated as the sum of right or left subitems 3.4, 3.5, 3.6, 3.7 and 3.8 (score range 0 to 20). The lateralized rigidity score was calculated as the sum of right or left subitems 3.3 (score range 0 to 8), respectively. When representing mean bradykinesia, rigidity and tremor in figures, we standardized scores on a 0 to 4 scale (as a single item) by dividing the mean by the number of summed items.

### Definition of contralateral side tremor development

Each patient had multiple assessments on the MDS-UPDRS over the 7-years span within the database. Since not all patients attended all the 7-year assessments, by convention, we defined a patient with a tremor fulfilling our contralateral TAS-criterium to become a TAS-patient for the rest of the observation period.

### On-assessments

All preceding methodological details refer to the participant in the off-state. We also considered the ON-state to assess the presence of contralateral tremor after taking the usual PD treatment. Because patients entering the PPMI were untreated and treatment was introduced after at least six months following enrollment according to participants’ clinical needs, not all subjects included in this analysis were assessed in the ON-state at every assessment. In **supplementary Table 1A and 1B** we have reported the number of participants with OFF and ON assessment at every follow up.

Because of this limitation, for the ON state we could only assess the cumulative number and percentage of participants with contralateral tremor on the total number of single participants that attended at least one follow up in the ON-state. More practically, 134 of 1S9 total participants with lateralized tremor had at least one ON-state assessment. Of these 134, 81 were right-dominant and S1 were left-dominant. Similarly to the OFF-state, in the ON-state we defined a patient with a tremor fulfilling our TAS-criteria to become a TAS-patient for the rest of the observation period.

### Statistical analysis

Descriptive categorical data is reported as numbers and percentages, while continuous scores are reported as mean and standard deviation (SD).

To investigate the development of TAS on the contralateral side, the Kaplan-Meier method was employed to generate the probability graph and associated descriptive tables.

To assess whether there was a significant progression over time of tremor, bradykinesia and rigidity lateralized scores (on the dominant and non-dominant sides), linear mixed-effect models fit by restricted maximum likelihood with random intercept were employed. Time (as a continuous variable), sex, age and disease duration at enrollment were used as predictors, as shown in the following equation:

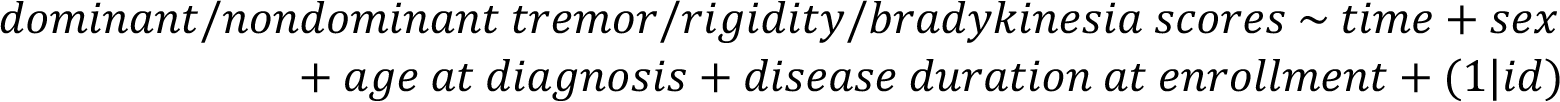

Statistical analyses were conducted in SPSS Statistics version 29 (IBM®, https://www.ibm.com/products/spss-statistics, RRID: SCR_002865).

### Data sharing

Data used in the preparation of this article was obtained on 2024-04-12 from the Parkinson’s Progressive Markers Initiative (PPMI) database (https://www.ppmi-info.org), RRID:SCR_006431. For up-to-date information on the study, visit www.ppmi-info.org. This analysis used data openly available from PPMI (Tier 1 Data). Codes generated to perform the analyses in this article are shared on Zenodo at the following DOI: doi.org/10.S281/zenodo.14892610.

## Results

### Cohort description

The PPMI cohort included in this study consists of 397 patients at baseline. We considered a maximum of 7 years follow-up for these. Demographic and basic clinical characteristics over the 7 years are reported in a previous study from our group,^3^ while in **Table 1** we summarize the main characteristics at baseline.

At baseline, out of the cohort of 397 patients, 1S9 participants (40.1% of the total cohort) showed TAS in the upper limbs, (**Table 2**). Of these, 95 showed a right dominant tremor and 64 a left-dominant tremor. The majority of these 1S9 participants showed rest tremor (138 subjects accounting for 8S.8%, 8S right-dominant, S3 left-dominant). 26 subjects also showed lateralized postural tremor (11 right-dominant, 1S left-dominant) and 27 subjects also showed lateralized kinetic tremor (10 right-dominant, 17 left-dominant). It should be noted that rest, postural and kinetic tremors may coexist in the same participants therefore the total number of the precedingly described tremor types (138+26+27=191) is not reflective of the single participants with lateralized tremor. Accordingly, in 26 participants a combination of rest, postural and/or kinetic tremor was present (**Table 3**). Notably, for all participants with more than one type of tremor, the different types of tremor were lateralized on the same side.

**Table 2.** Number of participants with no tremor, lateralized tremor, and bilateral tremor at baseline. This classification allows each patient to be classified into one and only one tremor group according to MDS-UPDRS tremor items 3.15, 3.16, 3.17 for rest, postural and kinetic tremor.

| **Severity of tremors**<br>**(worst tremor of the three tremor types - rest postural and kinetic)** | **Number of subjects**<br>**(total number: 397)** |
| --- | --- |
| **Patients with absent or slight tremor (both sides tremor <2)**<br>Consisting of: | **232 (58.4%)** |
| 1. patients with bilateral slight tremors (on both sides at least one tremor =1) | 52 (22.4% / 13.0%*) |
| 2. patients with lateralized slight tremors (one side at least one tremor =1, other side all tremors =0) | 119 (51.3% / 30.0%*) |
| 3. patients without tremor bilaterally (both sides =0) | 61 (26.3% / 15.4%*) |
| **Lateralized TAS (one side $\geq 2$, other side <2)\*** | **159 (40.1%)** |
| **Bilateral TAS (both sides $\geq 2$)\*** | **6 (1.6%)** |

*Abbreviations:* TAS, tremor amenable to surgery.
\* percentage based on patients with absent or slight tremor (232) / percentage based on the total (397) patients.

**Table 3.** Distribution of tremor types at baseline OFF-condition in participants with a lateralized tremor.

| **Lateralized TAS** | **Dominant tremor type** | **N of subjects** |
| --- | --- | --- |
| **Right sided tremor (95 subjects)** | Kinetic only | 6 |
|  | Postural only | 4 |
|  | Rest only | 76 |
|  | Two types | 7 (2 KT+RT, 5 PT+RT) |
|  | All three types | 2 |
| **Left sided tremor (64 subjects)** | Kinetic only | 6 |
|  | Postural only | 1 |
|  | Rest only | 40 |
|  | Two types | 13 (4 KT+PT, 3 KT+RT, 6 PT+RT) |
|  | All three types | 4 |

*Abbreviations.* TAS, tremor amenable to surgery.

### Emerging tremor on the contralateral side

Of the 1S9 patients who had unilateral TAS at baseline, cumulatively 2S.2% developed a TAS on the second side at 3 years, 30.8% at 5 years and 38.4% at 7 years. **Figure 1A** represents the probability function of developing contralateral TAS in the examined group estimated with the Kaplan-Meier method; **supplementary Table 2** shows the associated survival table. In 61 patients that developed contralateral tremor, the mean time to event is 3.23 years (median 3.0) and SD 2.09. Rest tremor affected the majority of these 1S9 participants with lateralized tremor and was by far the most common type of tremor (**Table 3**).

**Fig 1.**
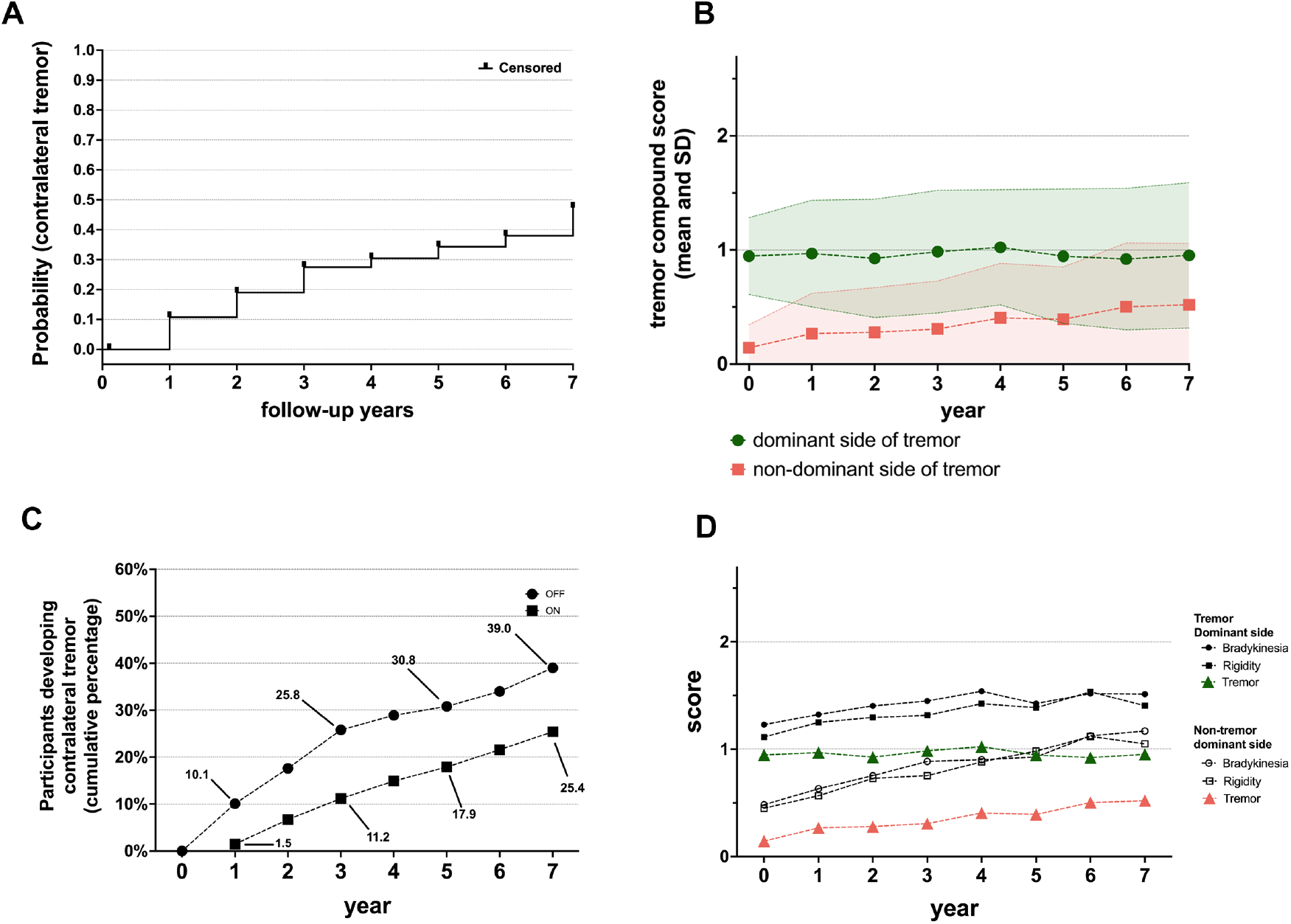
**(A)** represents the probability function for the development of contralateral TAS i.e. a score equal or greater than 2 in any of the tremor subitems of MDS-UPDRS items 3.16, 3.17 or 3.18) estimated with the Kaplan-Meier method. **(B)** represents the tremor severity (mean ± SD) of the OFF-state lateralized compound tremor score for postural, kinetic and rest tremor. Dominant versus non-dominant tremor sides were merged for all 159 patients. **(C)** shows the cumulative percentage of patients developing TAS over the seven years period comparing OFF-state (considering the initial 159 participants with lateralized tremor) and ON-state assessments (considering the subgroup of 134 participants with lateralized tremor that attended at least one ON-state assessment during follow up). (**D**) shows the mean (standardized 0 to 4) lateralized scores for bradykinesia, rigidity and tremor in the 159 lateralized TAS patients for each side separately at each available follow up visit. Linear mixed model showed that all compound scores worsen except for the tremor on the initially affected side.

The development of contralateral TAS was also examined in the ON-state. Due to the study design of the PPMI (participants initially unmedicated, off-assessment in clinic followed by ON-assessment if participant is treated), we were only able to assess how many participants cumulatively developed a TAS in ON-state. Of the 1S9 participants with baseline lateralized TAS, 134 (81 right-dominant and S1 were left-dominant) attended at least one follow up on-assessment. **Supplementary Table 1B** shows how many participants attended the ON state assessment at each follow up. **Figure 1C** represents the ON (and OFF) cumulative percentage of participants who developed contralateral TAS. 23.1% of 134 participants after 7 years showed a contralateral TAS, compared to 38.4% for the off-assessments.

Finally, we analyzed the progression of tremor, rigidity and bradykinesia compound scores separately for the dominant and non-dominant side (dominance was defined by the side with greater tremor at baseline) **(Fig. 1B** and **1D)**. Interestingly the compound tremor score increased only on the initially non-affected side but the rigidity and akinesia scores worsened significantly on both sides (**Table 4 and supplementary table 3**).

**Table 4.** Results of the linear mixed effects models to test the effect of time, sex, age at diagnosis, disease duration at enrollment on bradykinesia, rigidity and tremor scores on the dominant and non-dominant sides.

|  | Bradykinesia |  | Rigidity |  | Tremor |  |
| --- | --- | --- | --- | --- | --- | --- |
|  | Dominant side | Non dominant side | Dominant side | Non dominant side | Dominant side | Non dominant side |
| Follow-up time, estimate (SE) | 0.247<br>(0.058)*** | 0.513<br>(0.055)*** | 0.098<br>(0.026)*** | 0.183<br>(0.024)*** | -0.008<br>(0.036) | 0.205<br>(0.028)*** |

Note: The variable “Time” refers to follow-ups, once every year after baseline. The variable “Sex” was coded as 0 for females and 1 for males. Age and disease duration are considered at enrollment. Significance level:
\* p < 0.05; \*\*p < 0.01; \*\*\*p < 0.001

## Discussion

The lateralized progression of tremor in Parkinson’s disease has not been studied, although it is a clinically highly important information for invasive procedures, especially if initially directed to a single-hemisphere (unilateral deep brain stimulation or MR-guided focused ultrasound). This study shows that among the 1S9 patients in the PPMI cohort with significant, lateralized tremor (unilateral TAS), the contralateral side also developed TAS in 30% of cases at 5 years and 40% of cases at 7 years. These data are notable for patient counseling. Additionally, in this group we show that the dynamics of tremor worsening differ for the initially more affected side from the second affected side.

Interestingly, almost all 1S9 participants with lateralized TAS had a rest tremor, while only a minority had lateralized action tremor. Conversely, 232 participants had tremor scores <2 in both upper limbs. It is possible that action tremor is indeed less represented at this stage of the disease, although another possibility^7^is that action tremor amplitude continues to increase after 10 seconds of posturing, which is the temporal limit to observe postural tremor in the MDS-UPDRS item 3.15.

In our study, 87.7 % of patients, diagnosed at an early stage, had some kind of pathologic tremor manifestation at baseline.^2^ This is a high number and may reflect the fact that PD is suspected by non-specialists earliest when presenting with tremor, and subsequently these patients are more likely to be referred to specialized movement disorders clinics which recruited the PPMI-cohort. This cohort was diagnosed at an early stage of the disease of only 6 months. One of the characteristics of classic Parkinsonian tremor is that it starts mostly unilaterally. This is also true of other core symptoms of Parkinson’s disease. For the formerly used British Brain Bank criteria^8^ for the diagnosis of PD, the unilateral symptom dominance was even a core diagnostic criterion. In the entire PPMI group of 397 participants, 70% had a unilateral tremor with 40% severe enough to be classified as TAS.

A critical methodological issue of this study is the definition of TAS. We consider a tremor with an amplitude at the fingertips below 1-2 cm being less disturbing for the patient than a tremor above this amplitude. This is based mainly on clinical experience, because treatments are usually only considered successful if they reduce the tremor amplitude below this threshold.^9^ Consistent with this, tremor amplitudes above 1-2 cm are used as a threshold criterion for inclusion into MRgFUS studies.^6^ The amplitude at which surgery is no longer an option may vary by center and patient. Our chosen threshold is pragmatic also considering the experience-based definition of the scale anchors between ‘slight’ and ‘moderate’. In our view, it is not critical which tremor type (rest, posture, kinetic) is present in a particular patient, as all of them can be severely bothering.

To our knowledge, lateralization of tremor has not been analyzed in detail. While a different course of tremor progression on both sides may be clinically intuitive, the finding of a significant increase over time only in the contralateral side is somewhat unexpected. Indeed, in 1S9 participants with lateralized TAS, we found that mean tremor scores in this tremulous group do not further increase on the initially affected side during the first 7 years of disease. Conversely, bradykinesia and rigidity show significant increases on both sides. Of course, the lack of significant worsening of ipsilateral tremor average scores is influenced by the peculiar selection of participants in this analysis, which was tailored to understand the progression in PD patients with a very lateralized tremor onset. This finding may contribute to explain why earlier studies on tremor progression did not find a worsening.^10^ A recent study in a large cohort of early to mid-stage PD found that while overall bradykinesia and rigidity scores worsened over two years, rest tremor severity did not change significantly, while action tremors even decreased.^11^ The worsening of the overall tremor scores in the group is therefore mainly due to the worsening of the initially unaffected side developing tremor later. This was found despite similar worsening of bradykinesia and rigidity on both sides. The reason why tremor does not further increase from a certain time point on is not known. Different explanations come into play, two of them will be mentioned here: First, a simple scale effect, as the anchors of the MDS-UPDRS separate between tremors between below 1 cm, 1-3 cm, 3-10 cm and above 10 cm. The worsening could go in smaller steps and could only be detected with wearables. The amplitude of tremor depends also on physical factors like the size and weight of the hand and obviously the maximum amplitude is clearly limited.

Progression of PD symptoms has been a topic over the past 40 years. While early evidence suggested that patients with predominant tremor have a slower overall disease progression and longer time to death^12-14^ this has meanwhile largely be replaced by the better prognostic value of non-motor features.^1s^ Tremor in Parkinson’s disease is not related to a neuropathological lesion pattern^16^. Functional parameters of presynaptic striatal function like measures of dopamine synthesis and storage (Fluoro-Dopa-PET) or labelling of presynaptic transport proteins (FP-CIT, DatScan_SPECT) correlate nicely with bradykinesia and rigidity and even with the overall disease severity but not with tremor.^17^ The overall plateau effect of tremor scores, together with multiple other factors (e.g. non-dopaminergic neurotransmitter involvement), might explain some of these open questions.

A major limitation of this study is its limited follow-up time, but 7 years of follow-up obtained from such a consistently recruited cohort is almost unique information. In addition, the patients in this cohort were recruited after an average of 6 months of clinical symptoms and therefore the cohort is relatively homogeneous. Secondly, the PPMI-cohort may not be representative of the whole spectrum of early PD-manifestations as tremor-patients may have been overrepresented, but this does not affect the present study dealing exactly with this subgroup.

In conclusion, in this study we present data that may be clinically helpful for patients with unilateral tremor. Indeed, we have shown that 40% of patients have TAS after 7 years. Furthermore, we highlighted a lack of significant tremor progression in the initially-tremulous side, accompanied by a significant tremor score increase contralaterally. This finding contributes to explain the overall lack of tremor progression found in some previous studies.

## Data Availability

Data used in the preparation of this article was obtained on 2024-04-12 from the Parkinsons Progressive Markers Initiative (PPMI) database (https://www.ppmi-info.org), RRID:SCR_006431. For up-to-date information on the study, visit www.ppmi-info.org. This analysis used data openly available from PPMI (Tier 1 Data). Codes generated to perform the analyses in this article are shared on Zenodo at the following DOI: doi.org/10.5281/zenodo.14892610.

## Acknowledgement

We thank the colleagues of the PPMI-initiative for providing the data. Dr. C. Ferrer helped with insightful discussions on the topic.

## Authors’ Roles

Design: all: Execution: JC, NP, GD; Analysis: all; Writing: JC, NP, RH, GD; Editing of final version of the manuscript: All

## Financial Disclosures of all authors (for the preceding 12 months)

JP: none. Nicola Pavese has participated in advisory boards (Britannia, Boston Scientific, Benevolent AI, Roche, Abbvie), has received honoraria from Britannia, Abbvie, GE Healthcare, Boston Scientific, Teva Pharmaceuticals and grants from Independent Research Fund Denmark, Danish Parkinson’s disease Association, Parkinson’s UK, Center of Excellence in Neurodegeneration (CoEN) network award, GE Healthcare Grant, Multiple System Atrophy Trust, Weston Brain Institute, EU Joint Program Neurodegenerative Disease Research (JPND), EU Horizon 2020 research and innovation programme, Italian Ministry of Health. RC has received congress speech honoraria from AbbVie, Zambon, Bial, General Electric, EverPharma and grants from the Italian Ministry of Health, Italian Ministry of Research, Tuscany Region, European Joint Programme-Neuroscience Disease Research and the Fresco Foundation. GD has served as a consultant for Boston Scientific and Insightec. He receives royalties from Thieme publishers. He receives funding from the German Research Council (SFB 1261, T1) and private foundations. RH received funds for consultancy from Neurocrine Biosciences. RH received research grants from the MJ Fox Foundation, the Netherlands Brain Foundation, ParkinsonNL, and the Netherlands Organization for Health Research and Development.

## Supplementary material for

**Supplementary Table 1.** Number of participants with a lateralized tremor at baseline involved at each follow up visit with the off-state assessment (A) and on-state assessment (B). The number of total participants is also split by right

| **OFF** | Total participants, n | Right-dominant tremor participants, n | Left-dominant tremor participants, n |
| --- | --- | --- | --- |
| Baseline | 159 | 95 | 64 |
| Year 1 | 128 | 83 | 45 |
| Year 2 | 125 | 80 | 52 |
| Year 3 | 128 | 80 | 48 |
| Year 4 | 119 | 72 | 47 |
| Year 5 | 110 | 66 | 44 |
| Year 6 | 101 | 61 | 40 |
| Year 7 | 90 | 58 | 32 |

**B.**
| **ON** | Total participants, n | Right-dominant tremor participants, n | Left-dominant tremor participants, n |
| --- | --- | --- | --- |
| Baseline | 0 | 0 | 0 |
| Year 1 | 49 | 28 | 21 |
| Year 2 | 78 | 43 | 35 |
| Year 3 | 81 | 48 | 33 |
| Year 4 | 94 | 57 | 37 |
| Year 5 | 82 | 50 | 32 |
| Year 6 | 76 | 48 | 28 |
| Year 7 | 83 | 54 | 29 |

**Supplementary Table 2.**
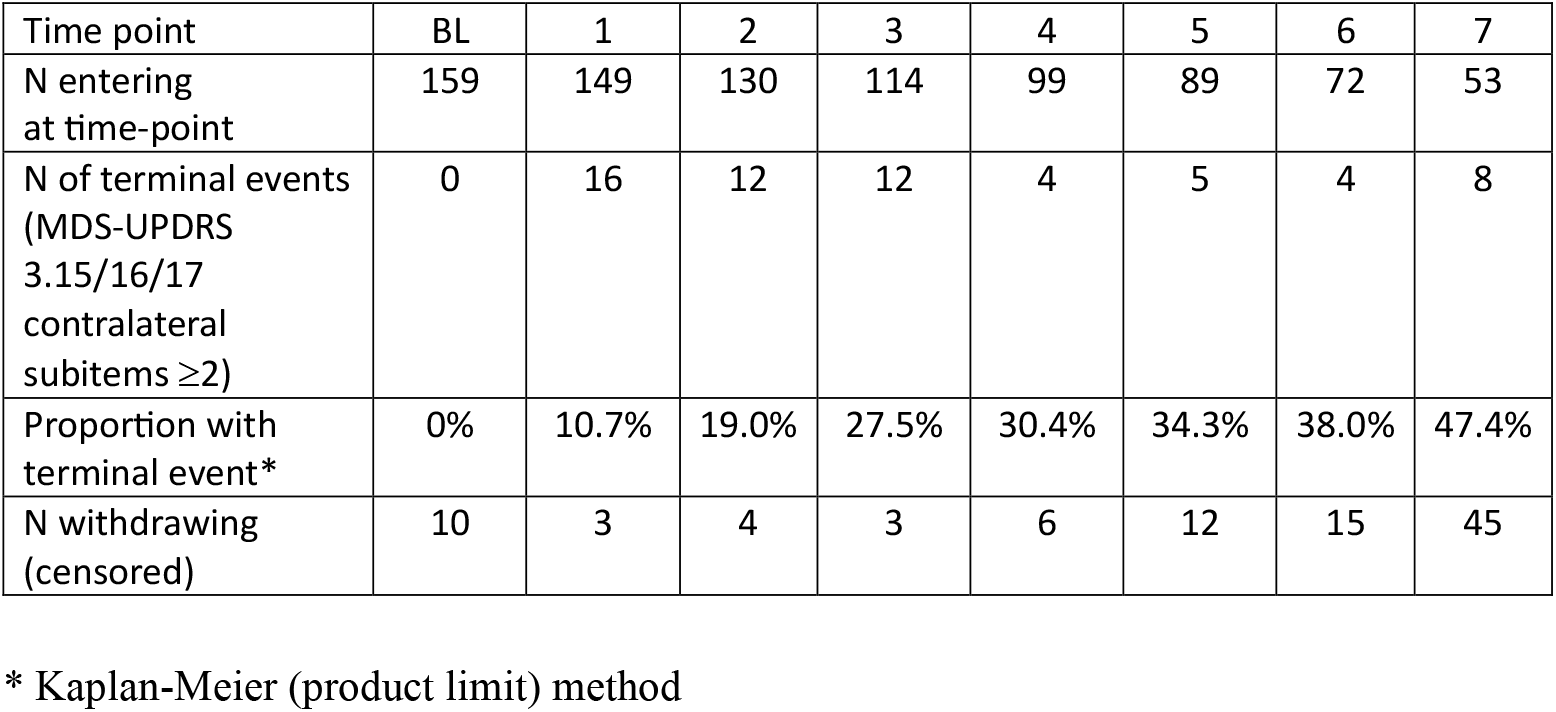
Survival table related to the development of contralateral upper limbs tremor score ≥ 2 in 159 participants with lateralized tremor at baseline.

**Supplementary table 3.**
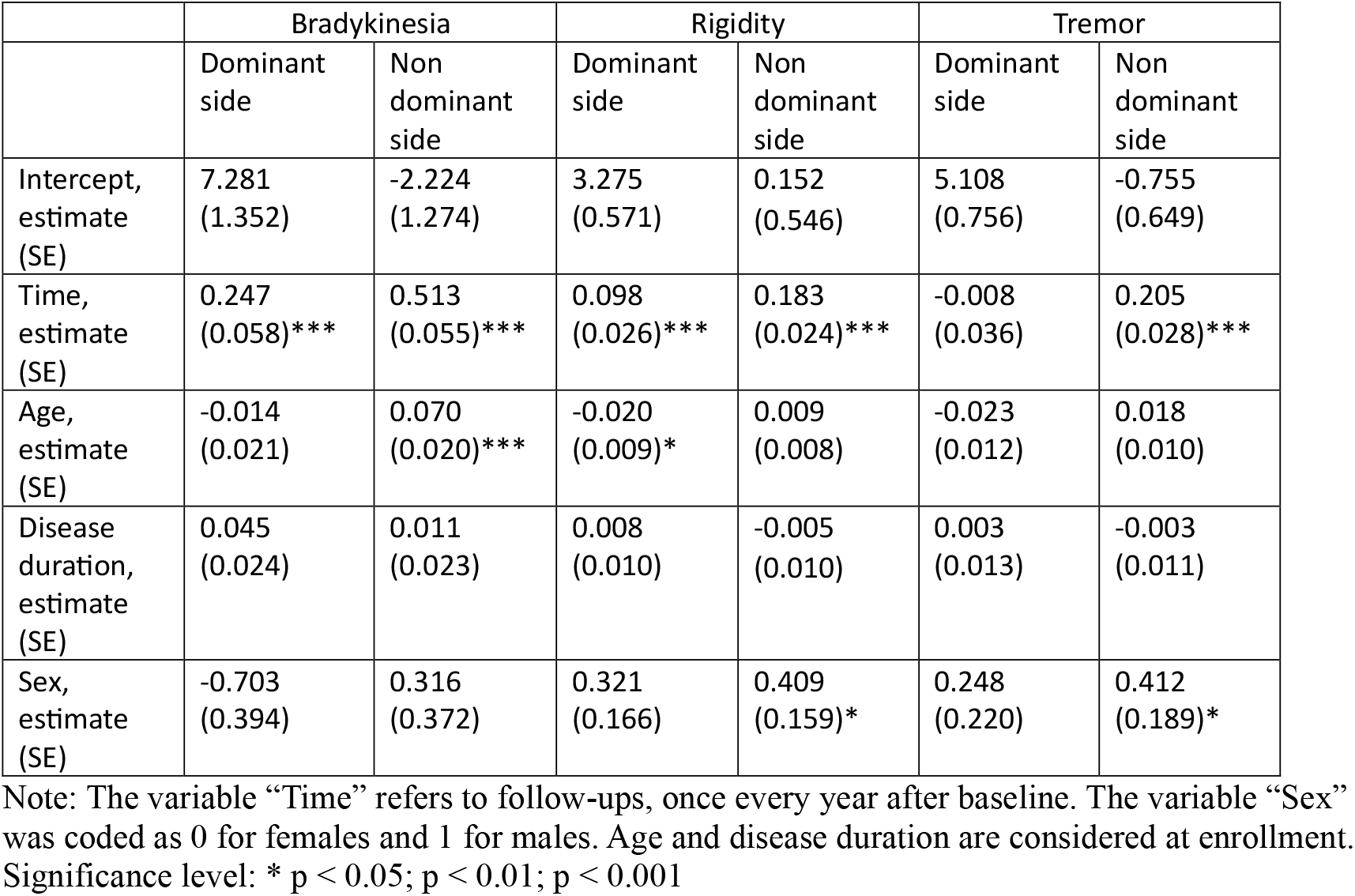
Results of the linear mixed models employed to assess the progression over time of bradykinesia, rigidity and tremor in the dominant and non-dominant sides (as defined by the side with greater tremor).

|  | Bradykinesia |  | Rigidity |  | Tremor |  |
| --- | --- | --- | --- | --- | --- | --- |
|  | Dominant side | Non dominant side | Dominant side | Non dominant side | Dominant side | Non dominant side |
| Intercept, estimate (SE) | 7.281 (1.352) | -2.224 (1.274) | 3.275 (0.571) | 0.152 (0.546) | 5.108 (0.756) | -0.755 (0.649) |
| Time, estimate (SE) | 0.247 (0.058)*** | 0.513 (0.055)*** | 0.098 (0.026)*** | 0.183 (0.024)*** | -0.008 (0.036) | 0.205 (0.028)*** |
| Age, estimate (SE) | -0.014 (0.021) | 0.070 (0.020)*** | -0.020 (0.009)* | 0.009 (0.008) | -0.023 (0.012) | 0.018 (0.010) |
| Disease duration, estimate (SE) | 0.045 (0.024) | 0.011 (0.023) | 0.008 (0.010) | -0.005 (0.010) | 0.003 (0.013) | -0.003 (0.011) |
| Sex, estimate (SE) | -0.703 (0.394) | 0.316 (0.372) | 0.321 (0.166) | 0.409 (0.159)* | 0.248 (0.220) | 0.412 (0.189)* |

Note: The variable “Time” refers to follow-ups, once every year after baseline. The variable “Sex” was coded as 0 for females and 1 for males. Age and disease duration are considered at enrollment. Significance level: \* $p < 0.05$ ; $p < 0.01$ ; $p < 0.001$

## References

1. Maetzler W, Liepelt I, Berg D. Progression of Parkinson’s disease in the clinical phase: potential markers. The Lancet Neurology 2009;8(12):1158–1171.

2. Postuma RB, Berg D, Stern M, et al. MDS clinical diagnostic criteria for Parkinson’s disease. Mov Disord 2015;30(12):1591–1601.

3. Pasquini J, Deuschl G, Pecori A, Salvadori S, Ceravolo R, Pavese N. The Clinical Profile of Tremor in Parkinson’s Disease. Movement disorders clinical practice 2023;10(10):1496–1506.

4. Pasquini J, Ceravolo R, Qamhawi Z, et al. Progression of tremor in early stages of Parkinson’s disease: a clinical and neuroimaging study. Brain 2018;141(3):811–821.

5. Marek K, Chowdhury S, Siderowf A, et al. The Parkinson’s progression markers initiative (PPMI) - establishing a PD biomarker cohort. Annals of clinical and translational neurology 2018;5(12):1460–1477.

6. Elias WJ, Lipsman N, Ondo WG, et al. A Randomized Trial of Focused Ultrasound Thalamotomy for Essential Tremor. N Engl J Med 2016;375(8):730–739.

7. Dirkx MF, Zach H, Bloem BR, Hallej M, Helmich RC. The nature of postural tremor in Parkinson disease. Neurology 2018;90(13):e1095–e1103.

8. Hughes AJ, Daniel SE, Kilford L, Lees AJ. Accuracy of clinical diagnosis of idiopathic Parkinson’s disease: a clinico-pathological study of 100 cases. J Neurol Neurosurg Psychiatry 1992;55(3):181–184.

9. Deuschl G, Raethjen J, Hellriegel H, Elble R. Treatment of patients with essential tremor. Lancet Neurol 2011;10(2):148–161.

10. Louis ED, Tang MX, Cote L, Alfaro B, Mejia H, Marder K. Progression of parkinsonian signs in Parkinson disease. Arch Neurol 1999;56(3):334–337.

11. van den Berg KRE, Johansson ME, Dirkx MF, Bloem BR, Helmich RC. Changes in Action Tremor in Parkinson’s Disease over Time: Clinical and Neuroimaging Correlates. Mov Disord 2024.

12. Roos RA, Jongen JC, van der Velde EA. Clinical course of patients with idiopathic Parkinson’s disease. Mov Disord 1996;11(3):236–242.

13. Zetusky WJ, Jankovic J, Pirozzolo FJ. The heterogeneity of Parkinson’s disease: clinical and prognostic implications. Neurology 1985;35(4):522–526.

14. Goetz CG, Stebbins GT, Blasucci LM. Differential progression of motor impairment in levodopa-treated Parkinson’s disease. Mov Disord 2000;15(3):479–484.

15. Fereshtehnejad SM, Romenets SR, Anang JB, Latreille V, Gagnon JF, Postuma RB. New Clinical Subtypes of Parkinson Disease and Their Longitudinal Progression: A Prospective Cohort Comparison With Other Phenotypes. JAMA Neurol 2015;72(8):863–873.

16. Paulus W, Jellinger K. The neuropathologic basis of different clinical subgroups of Parkinson’s disease. J Neuropathol Exp Neurol 1991;50(6):743–755.

17. Brooks DJ. PET studies on the function of dopamine in health and Parkinson’s disease. Ann N Y Acad Sci 2003;991:22–35.

